# Phase-Amplitude Coupling between Infraslow and High-Frequency Activities is a Potential Biomarker for Seizure Prediction

**DOI:** 10.1101/2020.11.07.20226258

**Authors:** Hiroaki Hashimoto, Hui Ming Khoo, Takufumi Yanagisawa, Naoki Tani, Satoru Oshino, Haruhiko Kishima, Masayuki Hirata

## Abstract

**IMPORTANCE:** This research describes a method to accurately predict the onset of epileptic seizures; this will help treat patients timely, prevent future seizures, and improve outcomes.

**OBJECTIVE:** We aimed to assess whether the phase-amplitude coupling (PAC) between infraslow activities (ISA) and high-frequency activities (HFA) increases before seizure onset.

**DESIGN AND SETTING:** This retrospective, single-center case series included patients admitted to the neurosurgery department at Osaka University Hospital in Suita, Osaka, from July 2018 to July 2019.

**PARTICIPANTS:** We enrolled seven patients with drug-resistant focal epilepsy who underwent intracranial electrode placement as part of a presurgical invasive electroencephalography study.

**MAIN OUTCOMES AND MEASURES:** We comparatively analyzed the ISA, HFA, and ISA-HFA PAC in the seizure onset zone (SOZ) or non-SOZ (nSOZ) in the interictal, preictal, and ictal states.

**RESULTS:** We recorded 15 seizures in seven patients [1 female (14%); mean (SD) age = 26 (12) years; age range, 15-47 years]. HFA and ISA were larger in the ictal states than in the interictal and preictal states. During seizures, the HFA and ISA of the SOZ were larger and earlier than those of nSOZ. In the preictal states, the ISA-HFA PAC was larger than that of the interictal states, and it began increasing at 93 seconds before the seizure onset (95% confidence interval: −116 – −71 s). There were no differences in the values and time of ISA-HFA PAC between both zones. Our phase-based analysis revealed differences between the SOZ- and nSOZ-PAC. In SOZ, the HFA amplitudes were tuned at the trough of the ISA oscillations, and in nSOZ, the HFA amplitudes were tuned at the peak of these oscillations. The receiver-operating characteristic curve showed that the ISA-HFA PAC of the SOZ showed the highest discrimination performance in the preictal and interictal states, with an area under the curve (AUC) of 0.926. However, ISA-HFA PAC was not suitable to differentiate between SOZ and nSOZ (interictal AUC = 0.555, preictal AUC = 0.691, and ictal AUC = 0.646).

**CONCLUSION AND RELEVANCE:** This study demonstrated the novel insight that ISA-HFA PAC increases before the onset of seizures, regardless of the seizure onset zone. Our findings indicate that ISA-HFA PAC is a potential biomarker for predicting the onset of seizures and may be valuable to physicians who routinely treat epileptic patients.

**Key Points:** *Question:* Is phase-amplitude coupling (PAC) between infraslow activities (ISA) and high-frequency activities (HFA) a useful biomarker for seizure prediction?

*Findings:* In this case series study on 15 focal-onset seizures in seven epileptic patients who underwent intracranial electrode placement, we found that a PAC of the ISA phase and HFA amplitude achieved significantly higher values in preictal states than in the interictal states, and ISA-HFA PAC of the seizure onset zone (SOZ) began increasing at 93 seconds before seizure onset (SO), while both HFA and ISA increased after SO. The receiver-operating characteristic curve showed that the ISA-HFA PAC of the SOZ showed the highest discrimination performance in the preictal and interictal states, with an area under the curve of 0.926.

*Meaning:* This study demonstrates that ISA-HFA PAC can differentiate between the preictal and interictal states of a seizure, indicating that it is a potential marker for seizure prediction.

## Introduction

Epileptic seizures are common neurological disorders. Timely detection of seizures is important for physicians to diagnose and quantitatively measure epilepsy. It was recently reported that seizures can be detected by applying deep learning methods^1^.

Infraslow activities (ISAs) and high-frequency activities (HFA) are key biomarkers of seizure detection^2-7^. They are both included in wideband electroencephalogram measurements and are measured by intracranial electroencephalogram (iEEG). ISAs, which are also referred to as direct current (DC) shifts^8^, are low-frequency components. Clinically, modern AC amplifiers with a time constant of 10 seconds allow filter-setting to record ISA recording (cutoff frequency of a high-pass filter is above 0.016 Hz)^5^. HFAs can be physiological^9^ or pathological^10^; epileptic HFAs are usually > 80 Hz^4^ and are pathological-type. High-frequency oscillations (HFOs), which are subgroups of HFAs^7^, are isolated oscillations that stand out from the background and can be further classified into ripples (80-250 Hz) and fast ripples (250-500 Hz)^4,11,12^.

The relationship between HFA and ISA has not yet been established. Ictal HFA and ISA are likely to occur in the same contacts^2,5^. Both occur earlier than conventional iEEG changes, but ictal ISA is observed more often and may occur earlier than HFA^3,5,13^. The ictal ISAs that precede ictal HFAs are called active DC shifts, whereas ictal ISAs that occur immediately after the onset of conventional ictal EEG patterns and HFAs are called passive DC shifts^5^. In interictal states, an ISA accompanied by an HFA could be a useful surrogate marker of the epileptogenic zone––this is called a “red slow”^5,14^.

Phase-amplitude coupling (PAC) is used to investigate the relationship between the low- and high-frequency bands on the EEG^15^. PAC analysis measures the degree of synchronization between the phases with low-frequency oscillation and high-frequency amplitude^16^. Ictal HFA amplitudes coupled with δ ^17,18^, θ^19,20^, and α^19^ phases, and β-HFA coupling are useful markers for seizure detection^21^. These pathological PACs are usually accompanied by ictal HFA, whereas physiological PACs that are involved in motor-related HFAs (high γ band) appear before the HFA increases^22,23^. Physiological PAC may induce a delay in physiological HFA; however, it is not known whether a preceding seizure-related PAC can induce a delay in ictal HFA or not. Moreover, a coupling between a frequency of 0.1 Hz and HFA has been previously reported^17^, but it is unclear whether a similar coupling between 0.016 Hz and HFA exists.

In our previous case report, we revealed that the PAC between ISA and HFA preceded the seizure onset (SO) and proposed that ISA-HFA PAC might be a useful marker for seizure prediction^24^. In this study, we hypothesized that ISA-HFA PAC might precede the SO and help us discriminate between interictal and preictal states; this may have implications for clinical use in seizure prediction. To assess this, we extracted ISA using a 0.016–1 Hz bandpass filter and HFA using an 80–250 Hz bandpass filter. We used the synchronization index (SI)^16^ to investigate PAC.

## Methods

### Study setting and participants

The retrospective study was performed at Osaka University Hospital in Suita, Osaka, Japan, from July 2018 to July 2019. The study design was approved by the Ethics Committee of Osaka University Hospital (approval no. 19193) and informed consent was obtained using the opt-out method on our center’s website. We included seven patients with drug-resistant focal epilepsy who underwent intracranial electrode placement as part of a presurgical invasive EEG study (Table 1).

**Table 1.** Clinical profile of the enrolled patients. Seizure numbers are serial numbers of seizures in each participant. SOC, seizure onset contact; MTLE, mesial temporal lobe epilepsy; FIAS, focal-impaired awareness seizure; FBTCS, focal-to-bilateral tonic-clonic seizure; PLE, parietal lobe epilepsy; OLE, occipital lobe epilepsy; FAS, focal aware seizure * Focal resection surgery was not performed because it was impossible to detect the seizure onset zone.

| PATIENT<br>NUMBER | SEX | AGE AT<br>SURGERY | LATERALITY | PATHOLOGY | SEIZURE<br>NUMBER | SEIZURE<br>TYPE | NUMBER<br>OF SOC |
| --- | --- | --- | --- | --- | --- | --- | --- |
| P1 | Male | 20's | Right | MTLE | S1 | FIAS,<br>FBTCS | 1 |
|  |  |  |  |  | S2 | FIAS,<br>FBTCS | 1 |
|  |  |  |  |  | S3 | FIAS,<br>FBTCS | 1 |
|  |  |  |  |  | S4 | FIAS,<br>FBTCS | 1 |
|  |  |  |  |  | S5 | FIAS,<br>FBTCS | 1 |
| P2 | Male | 40's | Right | PLE | S1 | FIAS,<br>FBTCS | 5 |
| P3 | Male | 20's | Left | OLE* | S1 | FIAS,<br>FBTCS | 2 |
| P4 | Female | 10's | Left | MTLE | S1 | FIAS,<br>FBTCS | 6 |
|  |  |  |  |  | S2 | FIAS,<br>FBTCS | 6 |
| P5 | Male | 10's | Right | OLE | S1 | FIAS,<br>FBTCS | 4 |
|  |  |  |  |  | S2 | FIAS,<br>FBTCS | 4 |
|  |  |  |  |  | S3 | FIAS,<br>FBTCS | 4 |
| P6 | Male | 30's | Left | MTLE | S1 | FAS | 3 |
| P7 | Male | 20's | Right | MTLE | S1 | FAS | 2 |
|  |  |  |  |  | S2 | FAS | 2 |

### Intracranial electrodes

To acquire iEEG data, we used a combination of subdural grids (10, 20, or 30 contacts), strips (four or six contacts), and depth electrodes (six contacts) (Unique Medical Co. Ltd., Tokyo, Japan) that were placed by conventional craniotomy (details shown in eMethod).

### Data acquisition and preprocessing

We acquired iEEG signals using a 128-channel digital EEG system (EEG 2000; Nihon Kohden Corporation, Tokyo, Japan) at a sampling rate of 1 kHz and a time constant of 10 seconds. The BESA Research 6.0 software (BESA GmbH, Grafelfing, Germany) preprocessed the raw signals using a high-cut filter at 333 Hz to prevent aliasing and a 60-Hz notch filter to eliminate the AC line artifact and exported the data as a text file. This text file containing iEEG signals was imported into MATLAB R2020a (MathWorks, Natick, MA, USA) and iEEG signals were digitally re-referenced to a common average of all electrode contacts in each patient.

We saved the iEEG data every 60 minutes; therefore, one text file contained one 60-minute signal. We applied a bandpass filter to the entire 60-minute data to prevent edge-effect artifacts.

### Seizure onset and contacts

The SO was determined by conventional visual inspection of iEEG signals^25^. The contacts that showed initial epileptic changes immediately after the SO were related to the seizure onset zone (SOZ) and those that showed no initial epileptic changes immediately after the SO were determined as non-SOZ (nSOZ) contacts; we randomly chose the contacts in nSOZ contacts such that we had a similar number of nSOZ and SOZ contacts (Table 1) (eFigure1).

The time before the SO was the preictal state and the time after the SO was the ictal state. We randomly extracted the time from the iEEG data without seizures such that the count of time values was ten times that of the SO; we defined this data as the interictal state.

### Infraslow activities and high-frequency activities

To extract ISA and HFA, we used a 0.016–1 Hz and 80–250 Hz bandpass filter, respectively. A bandpass device with a two-way, least-square, finite-impulse response filter (pop_eegfiltnew.m from the EEGLAB toolbox, https://sccn.ucsd.edu/eeglab/index.php) was applied to the iEEG signals. The oscillations of the 0.016–1 Hz signals were representative of ISA (eFigure2B and eFigure3B). We calculated the amplitude of HFA, which was an envelope of 80–250 Hz signals, in combination with the Hilbert transformation.

We combined a permutation test with a family-wise error (FWE)-corrected threshold to detect significant HFA changes. The oscillations under −1 mV or over +1 mV^2,13^ were defined as significant ISA changes (eMethod, eFigure2A, and eFigure4A, 4B).

### PAC analysis

An SI^16^ was used to measure the strength of coupling between the HFA amplitude and the ISA phase. Hilbert transformation was performed on the bandpass-filtered signals to obtain complex-valued analytic signals [Z(t)]. The amplitude [A(t)] and phase [φ(t)] were calculated from the complex-valued signals using Equation 1.

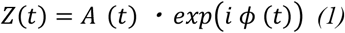

The ISA phase was calculated using the angle of the Hilbert transformation in the 0.016–1 Hz bandpass-filtered signal. The HFA amplitude was calculated using the squared magnitude of the Hilbert transformation in the 80–250 Hz bandpass filtered signal. Then, the phase of this amplitude was computed using the Hilbert transformation. The SI was calculated using Equation 2.

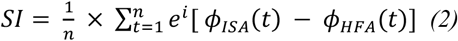

where “n” is the number of data points. Since SI is a complex number, we used the magnitude of SI (SIm) in our calculations. SIm varies between 0 and 1, with 0 indicating completely desynchronized phases and 1 indicating perfectly synchronized phases.

The preferred phase of synchronization (SIp) was calculated by the arctan (image [SI] / real [SI]). SIp varies between −180° and +180°.

### Phase-conditioned analysis

We calculated the mean vector and performed the Rayleigh test to evaluate the nonuniformity of SIp using the CircStat toolbox^26^. Next, to identify the ISA phase to which the HFA amplitude was coupled, we calculated the average oscillations of ISA and the normalized amplitude of HFA within each ISA phase bin of 30°: −180° – −150°, −150° – −120°, …, and 150° – 180°.

### Correlation analysis

Using all the implanted contacts from both SOZ and nSOZ, we calculated Pearson correlation coefficients between SIm and the normalized amplitude of HFA.

### Classification

We implemented a classification to distinguish between the three states using HFA, ISA, and PAC of SOZ or nSOZ. We also differentiated between SOZ and nSOZ was using the HFA, ISA, and PAC values of the interictal, preictal, and ictal states. The receiver-operating characteristic (ROC) curve and its area under the curve (AUC) were used to compare the performance of different classifiers.

### Statistical analysis

We compared SOZ with nSOZ within HFA, ISA, or PAC, and conducted paired comparisons of HFA with ISA, ISA with PAC, and HFA with PAC within SOZ or nSOZ using the Wilcoxon rank-sum test. The AUC was also compared using the Wilcoxon signed-rank test. The results were corrected using the Bonferroni correction for multiple comparisons.

## Results

Overall, we recorded 15 seizures, all of which were focal. We observed focal to bilateral tonic-clonic seizure (FBTCS) in 12 episodes (80%). There were 43 SOZ contacts (Table 1).

Representative seizures of S1 in P1 indicated that ISA and HFA changes occurred after the SO, whereas before the SO, there were no clear changes in them. However, SIm started to increase even a few minutes before the SO (eFigures 2 and 3).

### Profiles of HFA, ISA, and PAC

Within a 30-second time-window in the three states, we averaged and compared the HFA, ISA, and ISA-HFA PAC (SIm) of SOZ and nSOZ (Figure1A, 1B, and 1C, eMethod). In ictal-HFA and ictal-ISA, SOZ showed significantly higher values than nSOZ (corrected p < 0.001); however, there were no significant differences in ictal-PAC between SOZ and nSOZ. Both ictal-HFA and the ISA of SOZ achieved their maximum value, which was significantly higher than the interictal- and preictal-HFA and the ISA of SOZ (corrected p < 0.001). On the other hand, the ictal-PAC of SOZ was significantly lower than the preictal-PAC of SOZ (corrected p < 0.001). The preictal-PAC of SOZ achieved its maximum value and was significantly higher than the interictal-PAC of SOZ (corrected p < 0.001). Similarly, in the preictal stage, SOZ-ISA and nSOZ-PAC were significantly higher than those in the interictal stage; therefore, we believed that the PAC of SOZ and nSOZ and the ISA of SOZ might be potential biomarkers for seizure prediction.

Contrary to our expectation, the interictal- and preictal-HFA of SOZ were significantly lower than that of nSOZ (corrected p < 0.001 and = 0.005 for each).

### The time point when significant changes occurred

We compared the time points when HFA, ISA, and PAC of SOZ or nSOZ showed significant changes related to seizures (criteria for significant changes in eMethod and eFigure 4) (Figure 1D). In HFA and ISA, the SOZ occurred significantly earlier than nSOZ (corrected p < 0.001), whereas, in PAC, there were no significant differences between SOZ and nSOZ (corrected p = 0.31). Similarly, there were no significant differences between SOZ-HFA and SOZ-ISA (corrected p = 0.077); however, SOZ-PAC changed at a significantly earlier time (−93.82 s, 95% confidence interval: −116.40 – −71.23 s) than SOZ-HFA and SOZ-ISA (corrected p < 0.001). The average time of HFA and ISA was immediately after the occurrence of SO, whereas the average time of PAC was before the SO. We believe that these results also support the feasibility of ISA-HFA PAC for seizure prediction.

**Figure 1.**
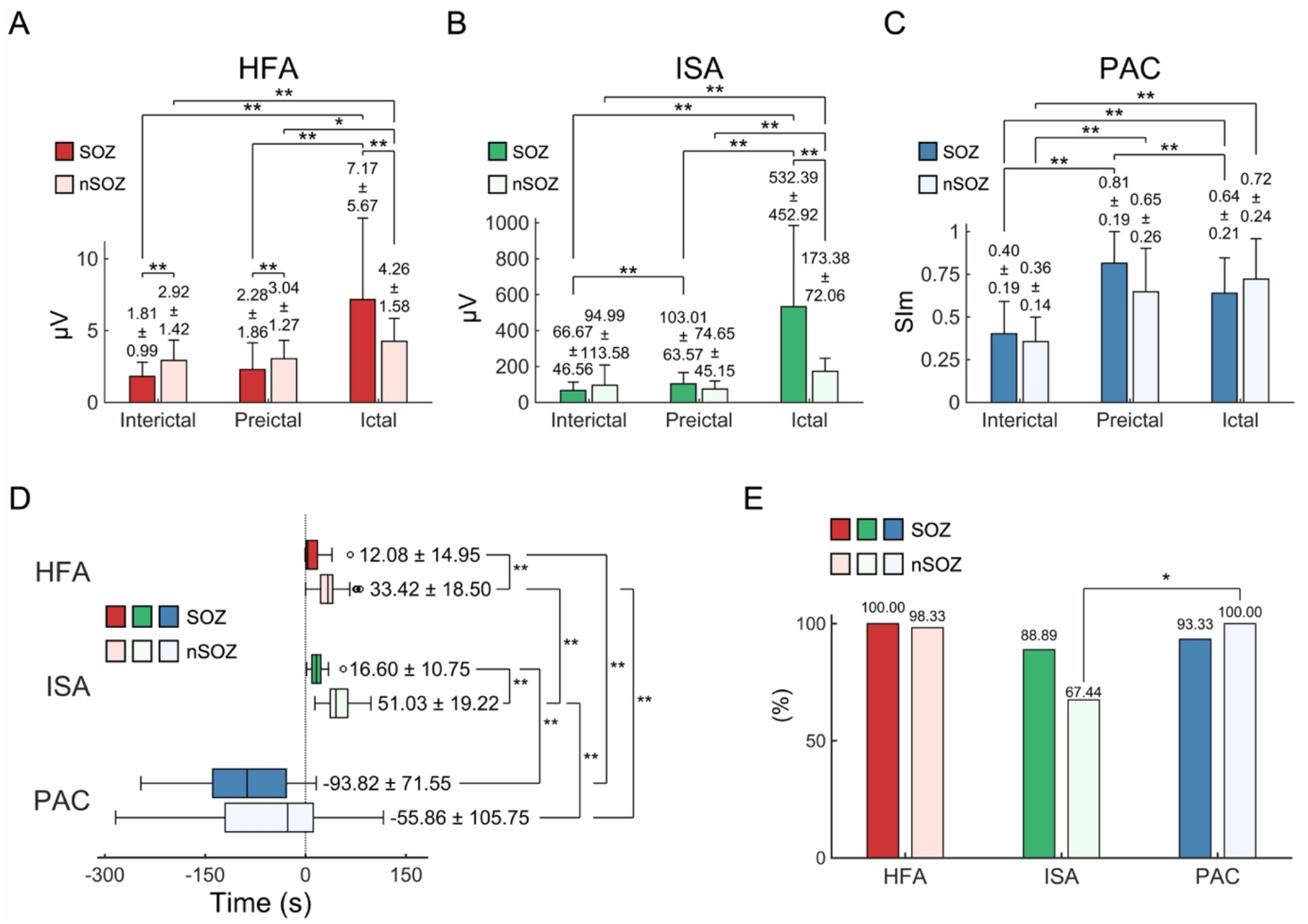
Profiles of HFA, ISA, and PAC. Paired results calculated from contacts placed in the SOZ and nSOZ are shown. (A) HFA amplitude of ictal-SOZ achieved its maximum value and was significantly higher than that of interictal-SOZ, preictal-SOZ, and ictal-nSOZ. (B) ISA oscillations of ictal-SOZ achieved their maximum values and were significantly higher than those of interictal-SOZ, preictal-SOZ, and ictal-nSOZ. (C) For ISA-HFA PAC, the SIm of preictal-SOZ achieved its maximum value and was significantly higher than that of interictal-SOZ and ictal-SOZ. Error bars in A, B, and C indicate the standard deviation (SD). (D) Time points at which we compared significant seizure-related changes, as shown in the box and whisker plots. For HFA and ISA, SOZ was significantly before nSOZ, and there were no significant differences between the times of SOZ and nSOZ-PAC. The average time of PAC was earlier than that of SO (0 seconds). (E) We observed the rate at which significant changes occurred. For HFA and PAC, almost all contacts of SOZ and nSOZ showed significant changes. For ISA, 89% of the SOZ contacts and 67% of the contacts of nSOZ showed significant changes. In panels A, B, C, and D, the average values and SD are indicated as figures. *p < 0.05, **p < 0.01, Wilcoxon rank-sum test, Bonferroni corrected.

### Rate at which significant changes were observed

Almost all the SOZ and nSOZ values showed significant changes in HFA and PAC. In ISA, 89% of SOZ and 67% of nSOZ showed significant changes. In the SOZ, there were no significant differences between HFA, ISA, and PAC (Figure 1E).

### Phase-based analysis

Figure 1 shows that in the ictal states, HFA- and ISA-SOZ had significantly higher values than nSOZ, and they changed earlier than nSOZ. However, PAC showed no significant differences between SOZ and nSOZ; therefore, we used phase-based analyses to investigate the differences between SOZ- and nSOZ-PAC.

Using a 30-second duration in the interictal or preictal states, we calculated the mean vectors of SIp in SOZ or nSOZ (Figure 2A); there were no similarities between SOZ and nSOZ in these two states. We observed significant nonuniformity only in the nSOZ-interictal state (p = 0.001, Rayleigh test).

**Figure 2.**
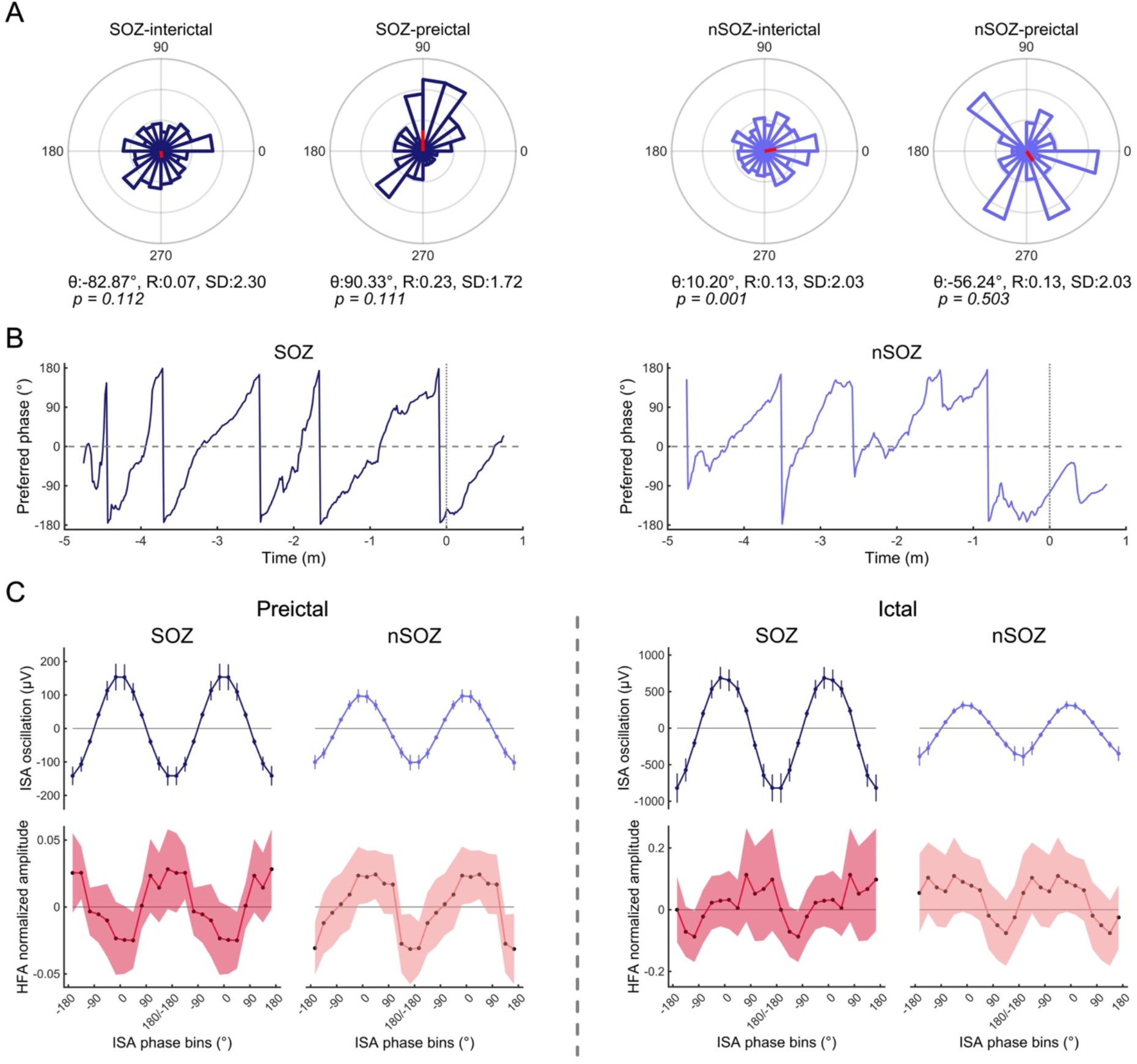
Phase-based analyses of PAC (A) We calculated average values of the SIp of SOZ or nSOZ in the interictal and preictal states. There were no similarities between the SOZ and nSOZ. The angle, length, and standard deviation of the mean vector are indicated as θ, R, and SD and p-values calculated by the Rayleigh test are shown. (B) We sequentially plotted the angles of the mean vector from −5 minutes to 1 minute. We observed continuous and periodic shapes in SOZ and discontinuous shapes in nSOZ. (C) The phase-tuning HFA-normalized amplitude was at its peak at the trough of the ISA phase and the bottom during the peak of the ISA phase in the preictal states; this trend was reversed in nSOZ. In the ictal states, these clear patterns became obscure. The error bars indicate 95% confidence intervals.

We sequentially plotted the angle (°) of mean vectors calculated from SIp in SOZ or nSOZ from −5 minutes to +1 minute around the SO (Figure 2B). In the SOZ, we documented continuous and periodic angle changes from −180° to +180°, while angle changes in nSOZ showed discontinuous and collapsed periodicity.

Figure 2C depicts the phases of ISA coupled with the amplitude of HFA, where “Preictal” indicates an average of 5 minutes (−5 to 0 minutes) and “Ictal” indicates an average of 1 minute (0 to 1 minute). In preictal-SOZ states, the HFA amplitude peaked at the trough of the ISA oscillation and reached its bottom-most value at the peak of the ISA oscillation, whereas we found an opposite trend in the nSOZ state––this was a contrasting result. These clear patterns were disrupted in the ictal state.

### Correlation between HFA and PAC

HFA increased in the ictal states, whereas PAC increased and displayed clear patterns in the preictal states rather than in the ictal states. After observing these contrasting results, we investigated the correlation between HFA and PAC.

There was no significant correlation between the preictal-HFA and preictal-PAC, and ictal-HFA and ictal-PAC; however, the correlation between the ictal-HFA and preictal-PAC was significantly positive (corrected p < 0.001) (Figure 3A). We calculated correlation coefficients (r) and their corrected p values in combination with sequential HFA and sequential PAC from −5 minutes to +2 minutes around the SO and displayed them as a matrix (Figure 3B). We observed an obvious positive correlation between ictal-HFA (after 0 minutes) and preictal-PAC (before 0 minutes). This demonstrates that contacts that indicated increased PAC before SO showed more of an increase in HFA after SO.

**Figure 3.**
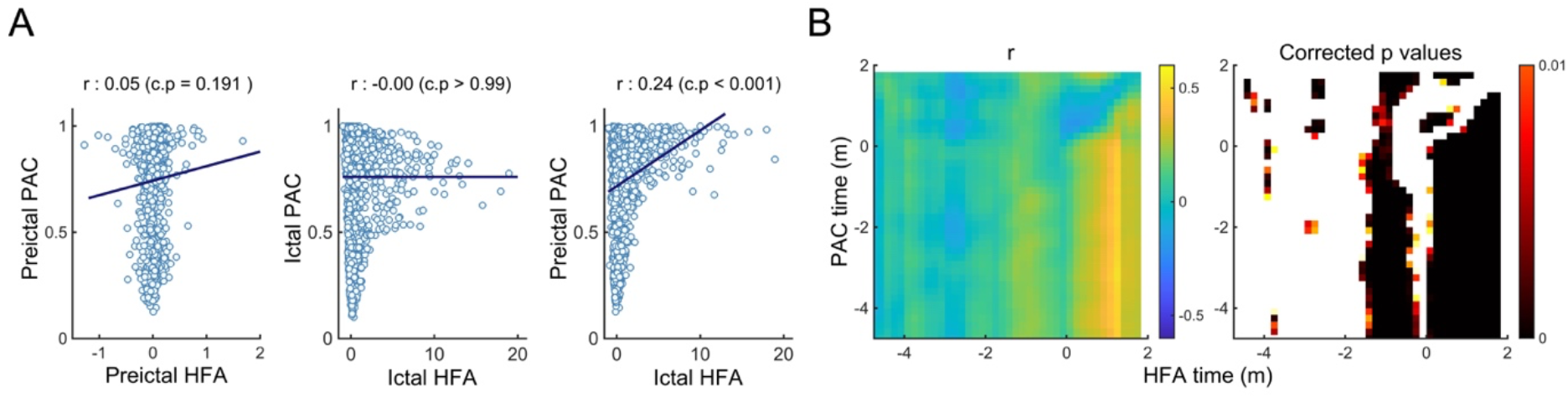
Correlation between HFA and PAC. (A) There was no significant correlation between HFA and PAC in the preictal or ictal state. However, ictal-HFA and preictal-PAC showed a significantly positive correlation (corrected p < 0.001). (B) Correlation coefficients and their corresponding significant p values (corrected p < 0.01) are shown for all combinations of sequential HFA and sequential PAC, from −5 minutes to +2 minutes around the SO (0 minutes). We observed a significant positive correlation in the range after the SO of HFA and before SO of PAC.

### Classification

To evaluate whether our method was accurately predicting seizures, we compared the preictal states with interictal states; to evaluate seizure detection, we compared the ictal states with interictal, and the ictal states with the preictal states (Figure 4A). After classifying the preictal and interictal states, we found that the AUC of SOZ-PAC was at its maximum and significantly higher than those of SOZ-ISA, SOZ-HFA, and nSOZ-PAC (corrected p < 0.001). SOZ-ISA showed the best performance in terms of seizure detection, better than SOZ-HFA, SOZ-PAC, and nSOZ-ISA (corrected p < 0.001, except for nSOZ-ISA in ictal vs. preictal; corrected p = 0.46).

**Figure 4.**
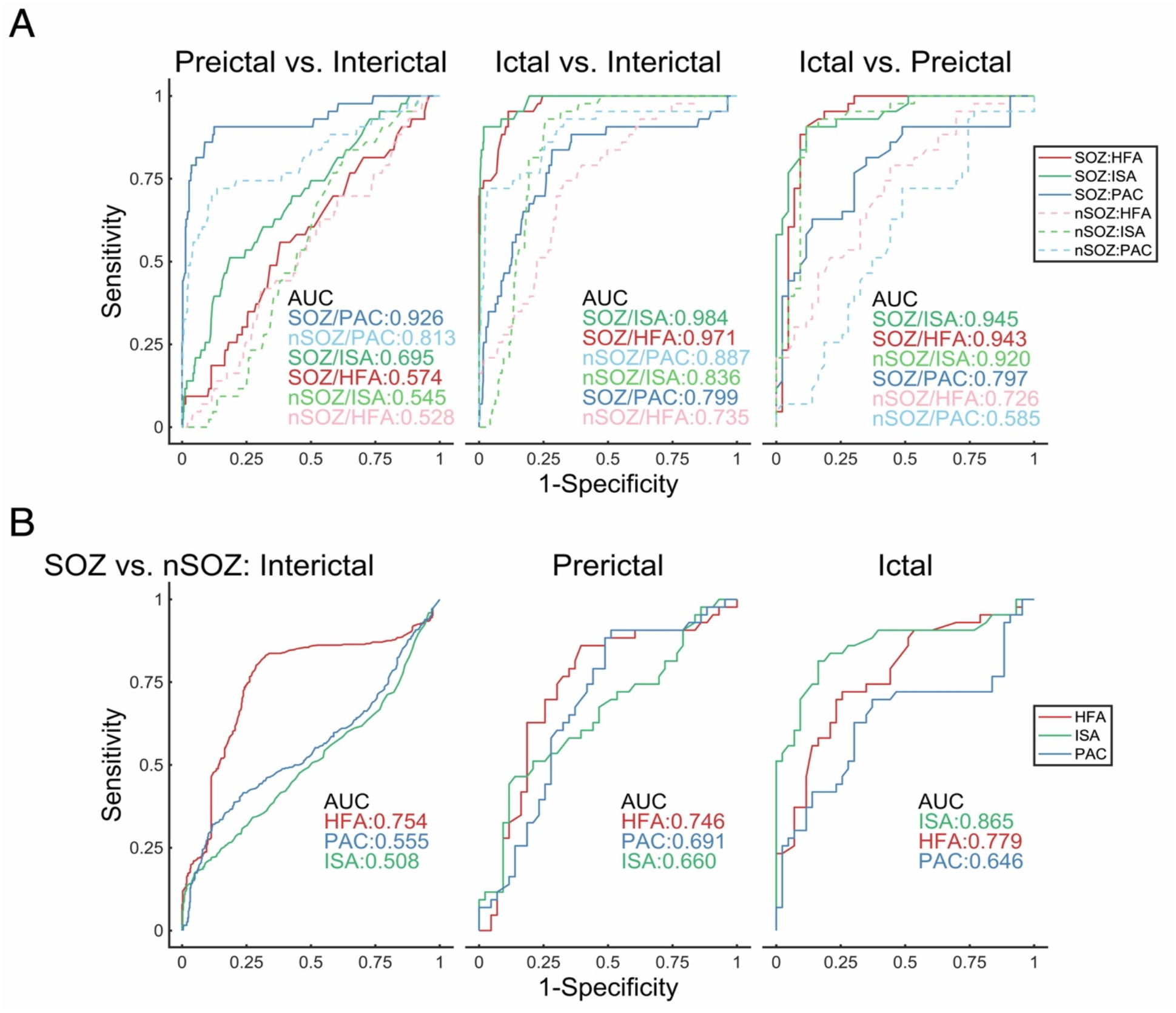
Average receiver operating characteristic curves of the results following stage classification. (A); Preictal vs. interictal, ictal vs. interictal, and ictal vs. preictal stages were classified using HFA, ISA, and PAC of SOZ or nSOZ. SOZ-PAC showed the highest performance between the preictal and interictal states, while SOZ-ISA had higher potential to separate the ictal and interictal, and ictal and preictal states. (B) On classifying SOZ and nSOZ using HFA, ISA, and PAC, we found that HFA had the maximum AUC in the interictal and preictal states. ISA performed best in the ictal states.

Finally, we classified SOZ and nSOZ using HFA, ISA, and PAC (Figure 4B). In the interictal states, PAC and ISA showed a chance level; however, HFA performed better than the others (corrected p < 0.001). Similarly, HFA showed the best performance in the preictal states (corrected p < 0.001). However, in the ictal states, ISA showed the best performance (corrected p < 0.001).

## Discussion

HFA and ISA are often observed during seizures^13,27-29^ and they are known to be useful biomarkers to detect SOZ^5,30,31^. In this study, SOZ achieved more ictal-HFA and ictal-ISA than nSOZ. The onset of ictal-ISA is typically earlier than that of the HFA^13,31^; however, we did not observe any differences between the onsets of SOZ-HFA and SOZ-ISA in this study. We defined the onset of ISA at the time point at > +1 mV or less −1 mV and the onset of ISA as the initial time of ISA reported in previous studies^13^. Therefore, our ISA onset time might be later than the values reported in previous studies. The percentage of SOZ-ISA in this study (88.89%) was concordant with that of a previous report (87%)^5^.

Our proposed PAC method brought new insight into the relationship between ISA and HFA. In this study, ISA-HFA PAC reached its maximum in the preictal state, began changing before SO, and showed no difference between SOZ and nSOZ. In the phase-based analyses, we observed differences between preictal-SOZ and preictal-nSOZ. The HFA amplitude in SOZ was tuned according to the trough of the ISA, which was concordant with the results of previous reports^19^; however, in nSOZ, the HFA amplitude was tuned at the peak of the ISA. These contrasting trends might be useful for differentiating between SOZ and nSOZ in preictal states.

During the seizure-dependent HFA increase, the PAC using a lower frequency (except for ISA) simultaneously increases^19,20,24^. However, in this study, PAC with ISA increased before the increase in HFA, which was concordant with the profile of physiological PAC^22,23^. Moreover, the PAC before SO was positively correlated with the later HFA; therefore, ISA-HFA PAC might play an essential role in inducing an HFA burst during or just before seizures.

Various features that are calculated using EEG have been proposed for seizure prediction^32^, and machine learning methods have been used to detect preictal states^33,34^. We observed a statistical significance between the interictal and preictal stages in SOZ-PAC, nSOZ-PAC, and SOZ-ISA (Figure 1); the AUC of preictal vs. interictal were big in this order (Figure 4). The AUC of SOZ-PAC was 0.926; therefore, SOZ-PAC could be a potential, novel biomarker for seizure prediction. Using ISA-HFA PAC, it is possible to achieve accurate and high-performing markers without complex algorithms and long training times. Accurate seizure prediction carries advantages for responsive neurostimulation with implantable devices^35,36^ and ultimately allows for early treatment and prevention of seizures.

Several features and algorithms have been used to detect seizures^32^, including the PAC of the β band^21^. Previously, ISA and HFA were simultaneously observed during seizures^5,13,31^, and our study added to the results by demonstrating that SOZ-ISA had a higher performance in differentiating between ictal states than SOZ-HFA. ISA and HFA are known to be useful for detecting SOZ^5,13,31^; we showed that in the ictal state, ISA performed better than HFA for SOZ differentiation.

Contrary to one expectation in our study, SOZ-HFA values were significantly lower than nSOZ-HFA in the interictal states. Reportedly, interictal-HFO is less correlated with the ictal onset zone^31^; therefore, interictal-HFA may be suppressed more in SOZ than in nSOZ. This surprising result further contributed to the interictal discrimination between SOZ and nSOZ.

This study has some limitations. Firstly, we obtained the result using mostly iEEG, and it is unclear whether the same results can be obtained using scalp EEG, which is the most commonly used method. Secondly, we evaluated only focal-onset seizures; therefore, it remains unclear whether generalized-onset seizures, such as absence or myoclonic seizures, would indicate the same results. Thirdly, we performed off-line analyses; however, in clinical situations, on-line analysis is fundamental for confirming the feasibility of seizure prediction using ISA-HFA PAC. We believe that further large-scale investigation is needed to strengthen our results.

## Data Availability

All data that were generated by or analyzed in this study are available from the corresponding authors upon reasonable request and after additional ethics approvals regarding the data provision to individual institutions.

## Author Contributions

H.H. conceived the study, collected the data, created the MATLAB program, analyzed the data, created all figures, and was primarily responsible for writing the manuscript. H.M.K., N.T., S.O., H.K., and M.H. performed the epileptic surgery. All authors clinically cared for and evaluated the patient. H.M.K., and T.Y. advised H.H. on scientific matters. H.K. and M.H. supervised this study. All authors have reviewed the manuscript.

## Supplementary Online Content

### eMethods

#### Intracranial electrodes

The diameter of each contact was 3 or 5 mm, and the inter-contact distances were 5, 7, or 10 mm for the grid and strip electrodes. The diameter was 1.5 mm and the inter-contact distance was 5 mm for the depth electrodes.

#### Topographies of HFA, ISA, and PAC

The 0.016–1 Hz bandpass-filtered signals for HFA, 80–250 Hz bandpass-filtered signals for ISA, and SIm signals for PAC were analyzed from 5 minutes before to 5 minutes after the SO. The power of the HFA was the square of the HFA amplitude (envelope) and the series was normalized by HFA power in the initial 1 minute (from -5 to -4 minutes before the SO). The HFA normalized power is shown as a topography (eFigure 3A). We defined the active seizure-related ISA as oscillations of the 0.016–1 Hz bandpass-filtered signals less than -1 mV or more than 1 mV; the active ISAs were displayed in grayscale topography (eFigure 3B). Statistically significant SIm values, acquired by a combination of a bootstrapped technique and familywise error (FWE)-corrected threshold (see below), were indicated in the topography (eFigure 4C).

#### Bootstrapped technique and FWE-corrected threshold

For the statistical assessment of SIm, we randomly shifted the phase-time series of the HFA amplitude and calculated the bootstrapped SIm (SImb) using the lower frequency phase. We repeated this procedure 1000 times to create the distribution of SImb^1^. The maximum values of SImb were stored at each iteration in its distribution. The values at 95% of the distribution of the maximum FWE-corrected threshold have been applied to the observed SIm to obtain the solution to multiple comparisons^2^. SIm values over the FWE-corrected threshold were statistically significant.

#### Profiles of HFA, ISA, and PAC (Figure 1A, 1B, and 1C)

For within-HFA comparisons, we used the amplitude of HFA signals, which were an envelope of 80–250 Hz bandpass-filtered signals. For within-ISA comparisons, the absolute values of oscillation of 0.016–1 Hz bandpass-filtered signals were used because the ISAs were slow-negative or slow-positive activities. For within-PAC comparisons, we used the SIm between the ISA phase and HFA amplitude.

We averaged values with a 30-second time window. The time interval of 30 seconds before the SO corresponded to preictal states, and that of 30 seconds after the SO corresponded to ictal states. The time of onset of the interictal states was randomly selected from iEEG data with no seizures.

#### Criteria for significant HFA, ISA, and PAC changes

To investigate the time when significant seizure-related changes occurred, we evaluated seizure-related signals from 5 minutes before to 2 minutes after the SO.

To detect significant changes in HFA, we used a permutation test^3^ to compare the initial 10-second data and the next 10-second data of HFA-normalized amplitude, which were acquired following normalization using the initial 10-second data. Each permutation test produced a set of differences between the initial 10-second data and the next sequential 10-second data. The maximum value of the differences from each permutation test was stored, and the values at 95% of the distribution of these maximum values were taken as the FWE-corrected threshold. The values above the FWE-corrected threshold were statistically significant^2^ and the time points when the HFA amplitude first crossed over the FWE-corrected threshold were defined as significant changes in HFA (eFigure 4A). However, if the HFA amplitude temporally crossed over the FWE-corrected threshold due to a single-spiking activity, we excluded it from the analysis.

For ISA, the time when iEEG signals first crossed under -1 mV or over +1 mV^4,5^ were defined as significant changes in ISA (eFigure 4B).

For PAC, we used significant SIm values, which were obtained in combination with the bootstrapped technique and FWE-corrected threshold. If the significant SIm formed a cluster, i.e., significant SIm was seen several times in 30-second time duration, we considered the first time point as the significant time of PAC changes (eFigure 4C).

**eFigure 1.**
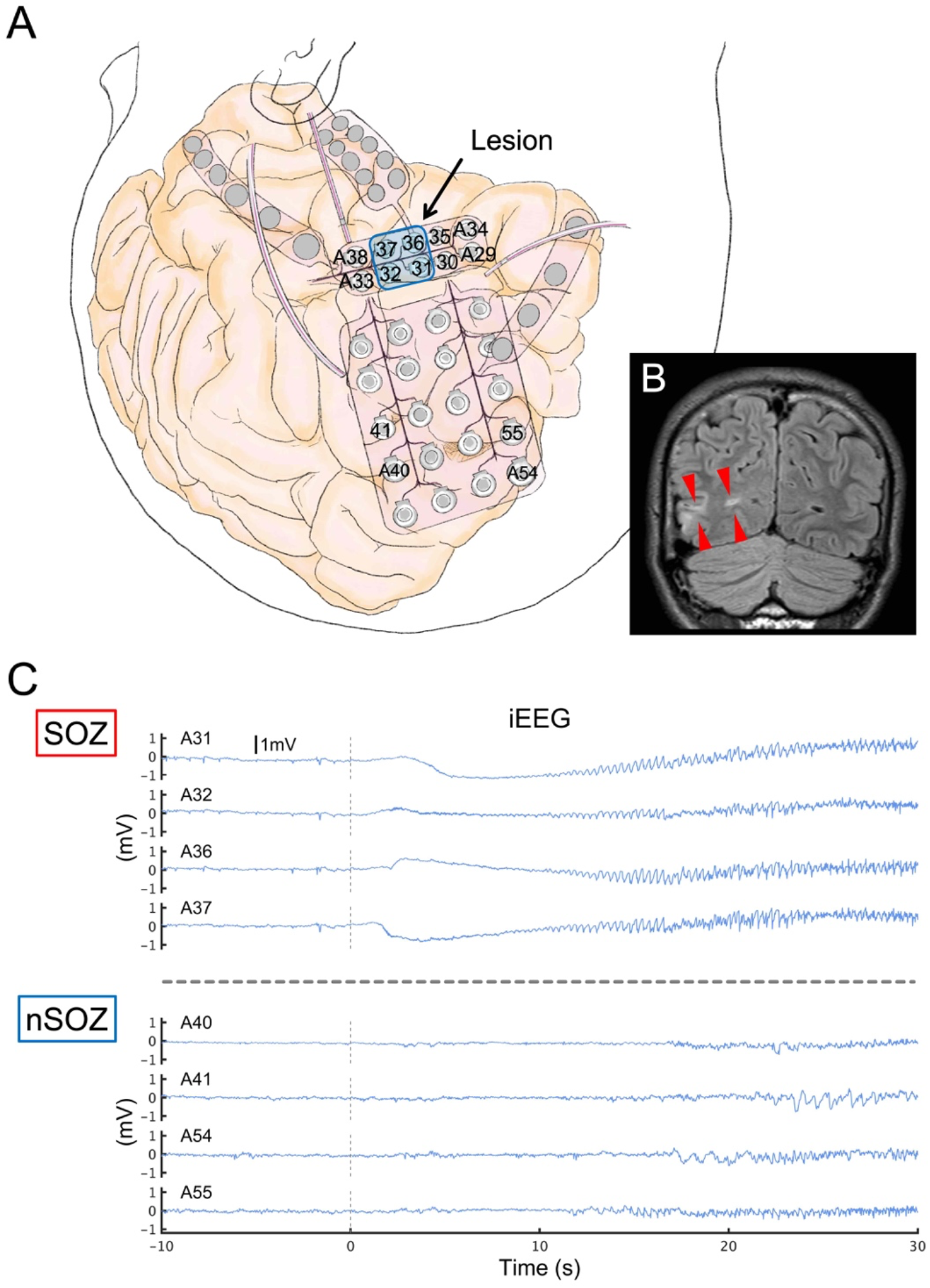
Contacts related to SOZ and nSOZ (P5-S3). (A) Illustration of implanted electrodes and the brain are displayed. The numbers correspond to the contacts’ number. (B) The high-intensity lesion in the right occipital lobe is shown on fluid-attenuated inversion recovery MRI (red wedge arrows) (B). This lesion is pointed by the arrow in the panel A. (C) SOZ contacts including A31, A32, A36, and A37 that were placed on the lesion showed initial infraslow activities immediately after the SO (0 s), followed by low-voltage fast waves. At that time, nSOZ contacts including A40, A41, A54, and A55 showed no epileptic changes. The number of SOZ and nSOZ contacts are same.

**eFigure 2.**
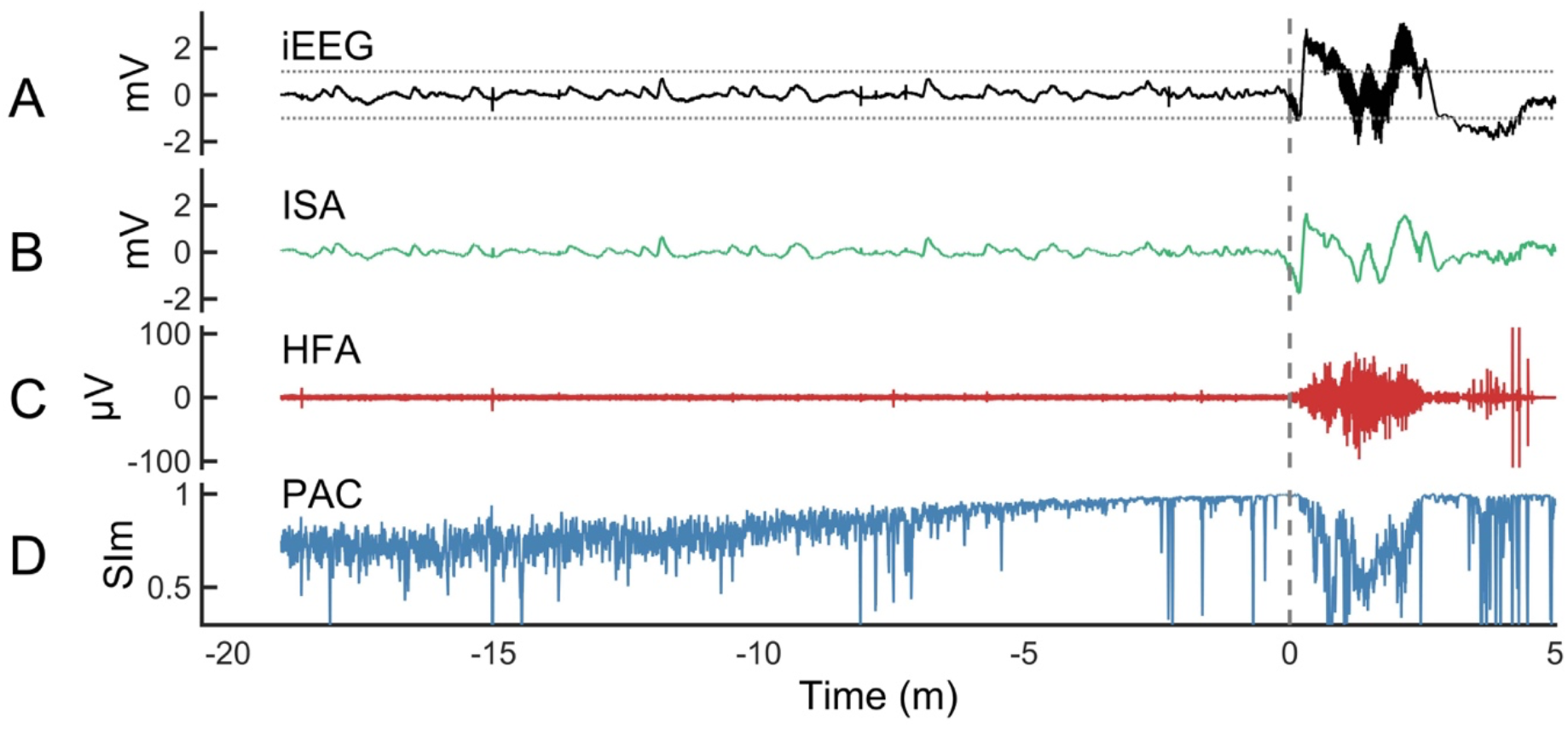
A representative seizure of S1 in P1. Representative multimodal signals related to seizure (S1 in P1 shown in Table 1) were calculated from one contact placed in the seizure onset zone (SOZ). From top to bottom column, raw iEEG signals in black (A), 0.016–1 Hz band-pass filtered signal as ISA in green (B), 80–250 Hz band-pass filtered signals as HFA in red (C), and SIm as ISA-HFA PAC in blue (D) are displayed. The 0 min corresponded to the SO. In iEEG signals, ± 1 mV were indicated as horizontal dotted lines, which were threshold for significant ISA related to seizures. After the SO, obvious activities over ± 1 mV were observed in iEEG signals, which were seizure-related ISA (A), and the same shape of signals was confirmed in ISA (B). Therefore, we knew that the 0.016–1 Hz band-pass filter enabled us to extract ISA activities. At the same time, an increase in HFA was observed, which was seizure-related HFA. However, before the SO, there were no obvious activities of either ISA or HFA. However, SIm started to increase from 15 min before the SO and reached a peak at the SO.

**eFigure 3.**
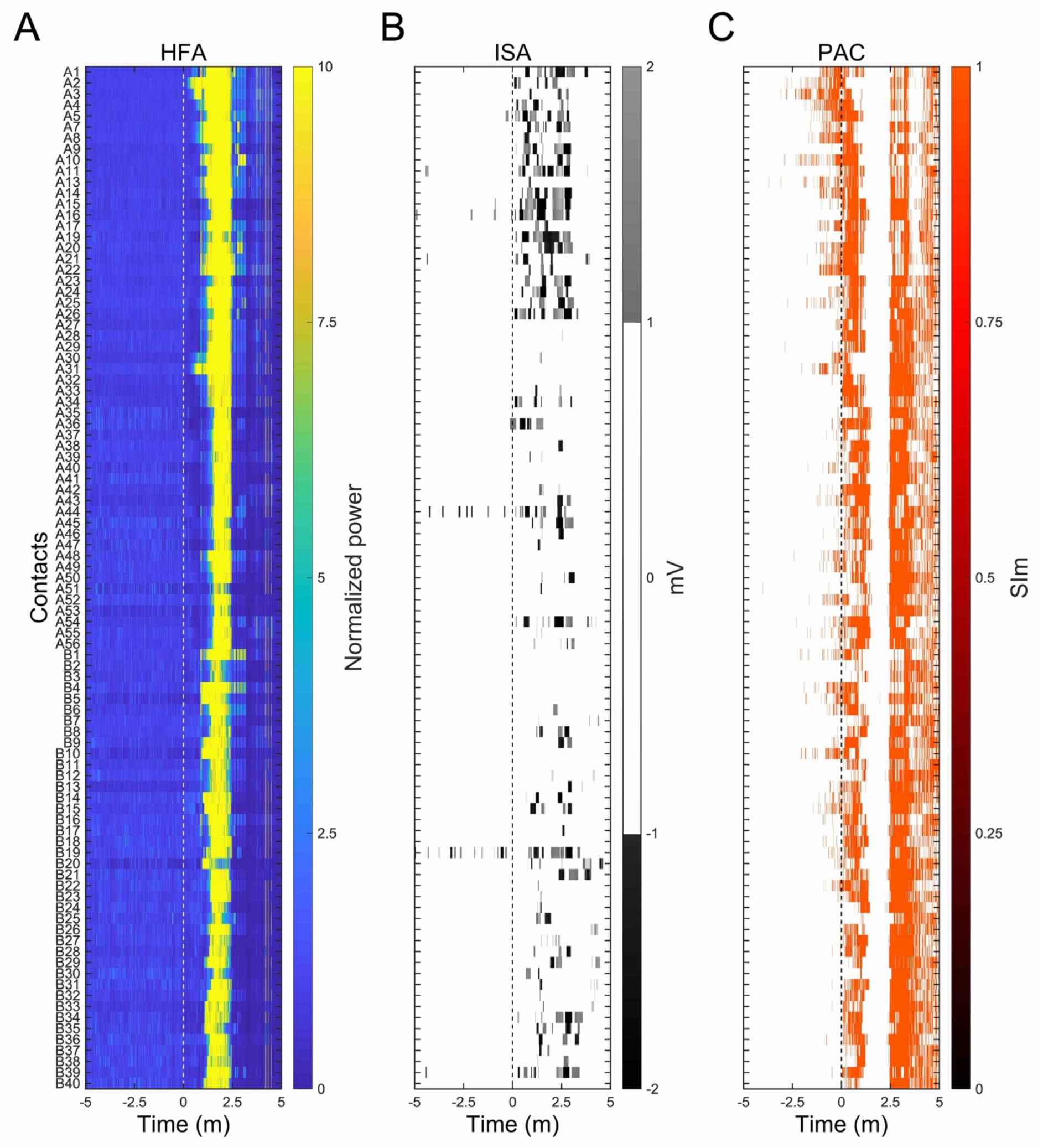
Topographies of ISA, HFA, and PAC of S1 in P1. The 0 minute-time point corresponded to the SO. All contacts of P1 were used; in eFigure 2 we only used the A2 contact that was placed in the SOZ. (A) The HFA-normalized power began increasing from the A2 contacts after SO and spreading to other contacts. (B) Active seizure-related ISAs, which were defined as more than 1 mV or less than -1 mV, are indicated on grayscale. There were obvious activities of both HFA and ISA after SO; however, no clear activities of either HFA or ISA were observed before SO. However, (C) we observed significant SIm with ISA-HFA, which was obtained using the FWE-corrected threshold that was applied to raw SIm, even before SO.

**eFigure 4.**
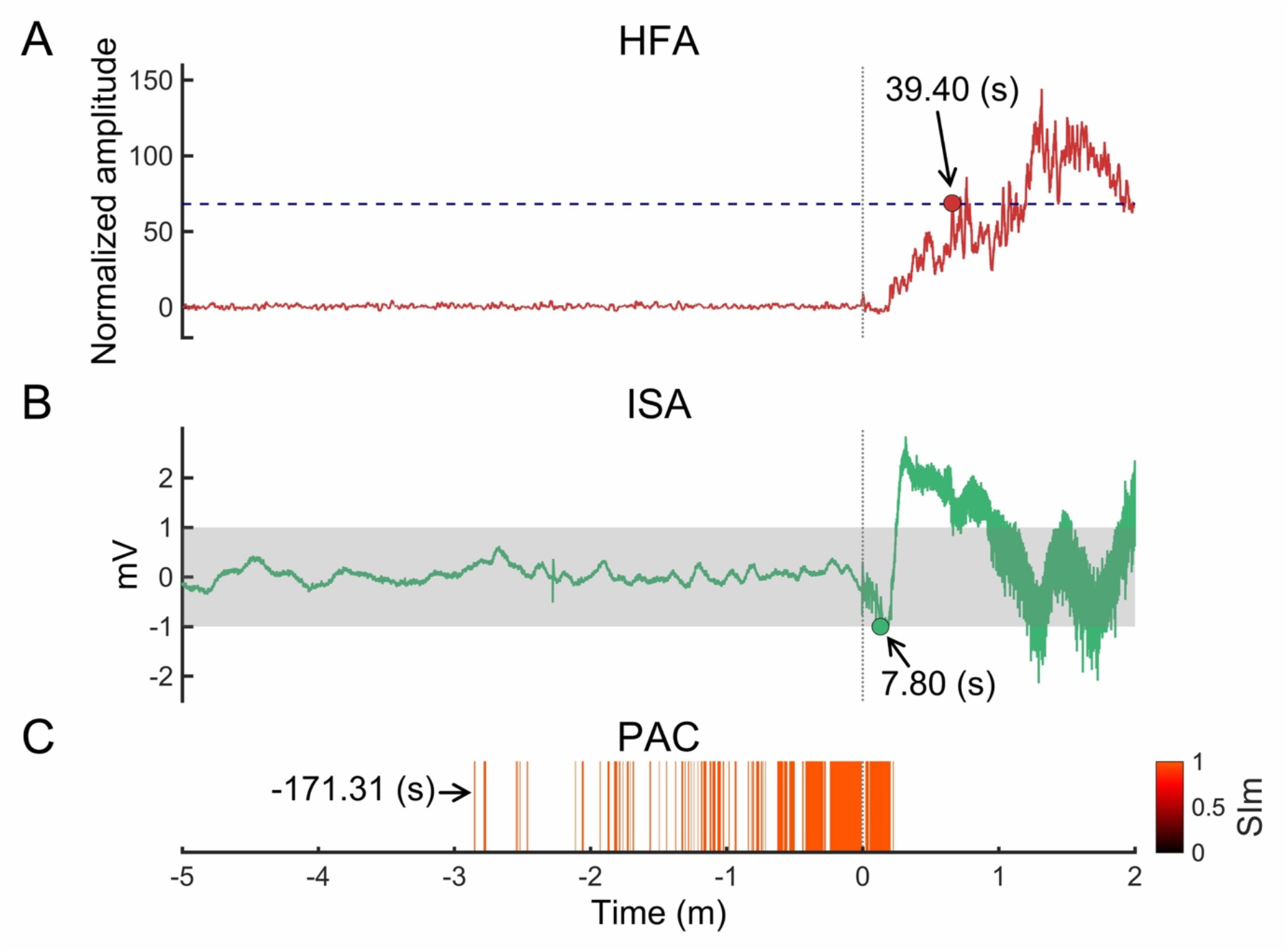
Period of significant change in S1 of P1. The seizure-related data from 5 min before to 2 min after the SO (0 min) of S1 in P1 are shown. (A) The HFA-normalized amplitude (red line) and FWE-corrected threshold (dashed blue line) are indicated. At 39.40 seconds, the HFA-normalized amplitude crossed over the FWE-corrected threshold for the first time, making this timepoint significant. (B) The iEEG signals (green lines) and range from -1 mV to +1 mV (gray mesh) are indicated. The time point of 7.80 seconds was the significant time of ISA change when the iEEG signal first crossed under -1 mV. (C) Statistically significant SIm appeared at -171.31 seconds, which was the significant time of PAC changes.

